# Environmental factors linked to reporting of active malaria foci in Thailand

**DOI:** 10.1101/2022.11.21.22281640

**Authors:** Preecha Prempree, Donal Bisanzio, Prayuth Sudathip, Jerdsuda Kanjanasuwan, Isabel Powell, Deyer Gopinath, Chalita Suttiwong, Niparueradee Pinyajeerapat, Ate Poortinga, David Sintasath, Jui A. Shah

**Affiliations:** Division of Vector Borne Diseases, Department of Disease Control, Ministry of Public Health, Nonthaburi, Thailand; Inform Asia: USAID’s Health Research Program, RTI International, Bangkok, Thailand; World Health Organization, Nonthaburi, Thailand; U.S. President’s Malaria Initiative, United States Agency for International Development (USAID), Regional Development Mission for Asia, Bangkok, Thailand; The SERVIR-Mekong Project, Asian Disaster Preparedness Center, Bangkok, Thailand

**Author notes:** These authors provided equal contribution to this study (Preecha Prempree, Donal Bisanzio).

**Keywords:** spatial analysis, environmental drivers, malaria elimination, disease modeling

## Abstract

**Background:** Thailand has made substantial progress toward malaria elimination, with 46 of the country’s 77 provinces declared malaria free as part of the subnational verification program. Nonetheless, these areas remain vulnerable to the reintroduction of malaria parasites and the reestablishment of indigenous transmission. As such, prevention of reestablishment (POR) planning is of increasing concern to ensure timely response to increasing cases. A thorough understanding of both risk of parasite importation and receptivity for transmission is essential for successful POR planning.

**Methods:** Routine geolocated case- and foci-level epidemiological and case-level demographic data were extracted from Thailand’s national malaria information system for all active foci from October 2012 to September 2020. A spatial analysis examined environmental and climate factors associated with remaining active foci. A logistic regression model collated surveillance data with remote sensing data to investigate associations with the probability of having reported an indigenous case within the previous year.

**Results:** Active foci are highly concentrated along international borders, particularly Thailand’s western border with Myanmar. Although there is heterogeneity in the habitats surrounding active foci, land covered by tropical forest and plantation was significantly higher for active foci than other foci. The regression results showed that tropical forest, plantations, forest disturbance, distance from international borders, historical foci classification, percentage of males, and percentage of short-term residents were associated with high probability to report indigenous cases.

**Conclusion:** These results confirm that Thailand’s emphasis on border areas and forest-going populations is well placed. The results suggest that environmental factors alone are not driving malaria transmission in Thailand; rather, other factors, including demographics and behaviors, may also be contributors. However, these factors are syndemic, so human activities in areas covered by tropical forests and plantations may result in malaria importation and potentially, local transmission, in foci that previously had been cleared. These factors should be addressed in POR planning.

## Background

Malaria remains a major public health threat across the world, with over 241 million cases and 627,000 deaths in 2020 [1]. The Greater Mekong Subregion (GMS), an area of Southeast Asia composed of Cambodia, Lao People’s Democratic Republic (PDR), Myanmar, Thailand, China’s Yunnan Province, and Vietnam, is now an area of low malaria incidence but high population connectivity [2]. To accelerate progress in the region, GMS countries have jointly committed to eliminating *Plasmodium falciparum* (*P. falciparum*) by 2025 and to eliminating all forms of human malaria by 2030, following guidelines set in the World Health Organization’s (WHO) Strategy for Malaria Elimination in the Greater Mekong Subregion (2015-2030) [3].

Thailand also established ambitious, yet attainable, national goals to interrupt malaria transmission, reduce active foci, and achieve nationwide malaria-free status by 2024 [4, 5]. The country’s keystone 1-3-7 surveillance strategy, adapted from China, requires that all malaria cases be reported within 1 day of detection and investigated within 3 days, and that appropriate focus-level response is completed within 7 days [6, 7]. Each case and focus are classified according to Thailand’s detailed protocols, and public health interventions are responsively tailored. Foci are categorized as active (A1), residual (A2), cleared but receptive (B1), or cleared but not receptive (B2), as detailed in ***Table 1*** [5]. In the initial 5 years of the 1-3-7 strategy’s implementation, malaria incidence decreased by 81%, from 14,954 cases in fiscal year (FY) 2017 to 2,835 in FY2021, and nearly all annual targets are being met [6]. Additionally, 46 of the country’s 77 provinces have now been declared malaria free as part of its subnational verification program.

**Table 1.** Focus classification in Thailand.

| Focus classification | Pre-2016 definition | Current definition |
| --- | --- | --- |
| A1 | Perennial transmission village or hamlet where indigenous cases are reported at least 6 months out of the year | *Active focus*: Reported indigenous transmission in the current year |
| A2 | Periodic transmission area village or hamlet where indigenous cases are reported fewer than 6 months out of the year | *Residual non-active focus*: No indigenous cases in the current year but with indigenous cases in the previous 3 years |
| B1 | High-risk village or hamlet without transmission for a minimum of 3 years, but adult vectors or larvae are present or conditions are favorable for breeding | *Cleared focus but receptive*: No indigenous transmission in at least 3 years, but suitable environment for vector *Anopheles spp.* mosquitoes |
| B2 | Low-risk village or hamlet without transmission for a minimum of 3 years, and no presence of adult vectors or larvae and unfavorable conditions for breeding | *Cleared focus but not receptive*: No indigenous transmission in at least 3 years, but unsuitable environment for vector *Anopheles spp.* mosquitoes |

Despite this progress, challenges to malaria elimination persist, including adequately addressing *Plasmodium vivax* (*P. vivax*) infections, heterogeneous epidemiology, and high population mobility [8, 9, 10]. In FY2013, 43.7% of malaria infections were due to *P. falciparum*, but by FY2020, the parasite only accounted for 5.7% of all infections. *P. vivax* has become the predominant species, causing 91.6% of malaria infections in 2020 [11, 12]. *P. vivax* infections can stay latent for a long time, are less likely to be symptomatic, and require a long 14-day treatment course of primaquine in Thailand as radical cure to prevent relapse, thereby challenging malaria elimination. The spatial distribution of malaria is highly heterogeneous across the country [13, 14]. Cases are clustered in difficult-to-reach populations and areas, often among male workers who travel along Thailand’s international borders [15]. Thai nationals still comprise a majority of malaria cases (71.3% in FY 2020); however, there is a disproportionately high malaria incidence among mobile and migrant communities [11]. As indigenous transmission decreases in Thailand, imported parasites and clustered infections in mobile populations have become an increasing concern. Imported cases can trigger local transmission in susceptible areas [16, 17, 18]. Simultaneously, Thailand’s subnational verification program has expanded the number of malaria-free areas. Although this designation is hard earned, these areas remain vulnerable to the importation of malaria parasites and the reestablishment of indigenous transmission. Both international and domestic travel may increase susceptibility in malaria-free areas or threaten progress toward Thailand’s malaria elimination goal.

As such, Thailand’s Division of Vector Borne Disease (DVBD) in the Ministry of Public Health (MOPH) has accelerated its provincial-level prevention of reestablishment (POR) planning to ensure timely response to increasing cases. The draft POR stratification incorporates both risk of parasite importation and receptivity for transmission, building on Thailand’s long-standing focus classification. The classification considers the presence of both mosquito vectors and habitats conducive to vectors, such as forests or mature rubber plantations, based on available entomological surveillance data. To further progress toward malaria elimination, however, focused efforts must be made that draw upon all available sources of information.

This study examined environmental, climate, and demographic factors associated with remaining active foci in Thailand. A logistic regression model collated both routine case- and foci-level surveillance data with remote sensing data to investigate associations of these factors with the probability of having reported an indigenous case within the previous year. The results are intended to contribute to understanding what factors are fueling current malaria transmission and potential future reintroduction, thereby enhancing the current POR risk stratification strategy and increasing the DVBD’s ability to disrupt transmission.

## Methods

### Malaria data

Routine geolocated case- and foci-level epidemiological and case-level demographic data were extracted from the national malaria information system (MIS). In Thailand, any focus that has recorded an indigenous case in the previous one year is classified as A1 (based on an annual focus classification cycle) and tracked as an “active focus” [5]. The analysis included all active foci in which at least one parasitologically confirmed (by microscopy or rapid diagnostic test) case from public health and non-governmental community facilities was reported over an 8-year period from October 2012 to September 2020, representing FY2013 to FY2020. Focus classifications were standardized according to the classification criteria adopted after 2015 to ensure consistency in definitions and to facilitate comparisons across the study period. Independent case-level variables for the regression analysis were drawn from reported cases from FY2016 to FY2020 (to match the timeline of available environmental data) with full demographics, travel history, and parasite species data. The study utilized fiscal years (October to September) because Thailand’s malaria program and database are based on fiscal year targets, and FY2020 was chosen as an endline to align with the availability of environmental data.

### Land use data

Environmental features around malaria foci were estimated using the Global Land Cover product with Fine Classification System at 30 m (GLC_FCS30-2020) land cover classification map obtained from Liu et al. (2020) [19]. The GLC_FCS30-2020 had a resolution of 30 m and higher granularity of land cover classification compared to other land cover products. Using the land cover map, we calculated the percentage of territory covered by crops, mixed forests, tropical forest, plantation, permanent and seasonal water bodies, built areas, and grasslands in a 1-km radius around each malaria focus. These categories are defined in detail elsewhere [20], but selected descriptions relevant to Thailand are listed below:

- *Cropland* includes herbaceous and shrubby crops such as cereals, oils seeds, vegetables, root crops and forages but excludes tea, coffee, and rice.
- *Plantation* includes perennial crops of >5 m such as rubber, palm oil, cashew, and coconut.
- *Tropical forests* have >60% canopy cover from trees at least 5 m tall, and the dominant tree species are evergreen broadleaf
- *Mixed forests* have >60% canopy cover from trees at least 5 m tall; forest is considered mix because no single forest type makes up >60% of the total tree cover.

The landcover calculation was performed using the open-source Geographic Information System Geographic Resources Analysis Support System (GIS GRASS) [21].

### Rainfall data

Global precipitation estimates were downloaded from the Global Precipitation Climatology Centre (GPCC) V7 [22], which considers the largest number of contributing ground observations [23]. For this study, GPCC V7 monthly precipitation data at 1-degree spatial resolution from 2016 to 2020 were obtained from the Deutscher Wetterdienst (DWD, https://opendata.dwd.de/) for all malaria foci.

### Forest disturbance data

Because deforestation in Thailand has been associated with malaria transmission [16], we included information about forest disturbance in the study analysis. Latest available raster data about forest disturbance on a global scale from 2015 to 2019 were obtained from the Global Forest Change website (http://earthenginepartners.appspot.com/science-2013-global-forest) [24]. The values reported in the data create a disturbance index indicating the level of disturbance on a scale from zero (no disturbance) to 17 (highest level of disturbance) at a 25-m resolution. We calculated the mean annual disturbance index in a 1-km radius around each malaria focus.

### Statistical analysis

Comparisons of percentage of territory covered by environmental features among the four foci classes were performed using the Kruskal-Wallis test. A logistic regression model was built to investigate the associations of environmental and demographic factors, with the probability of a focus to report an indigenous malaria case and thus be classified as an active focus (A1). This project employed a Structural Additive Regression (STAR) model [25]. This model’s structure allows the inclusion of both linear and non-linear predictor effects, increasing the model’s flexibility. The STAR model can also include a spatial spline factor to account for spatial autocorrelation of the data. The model’s full formula is below:

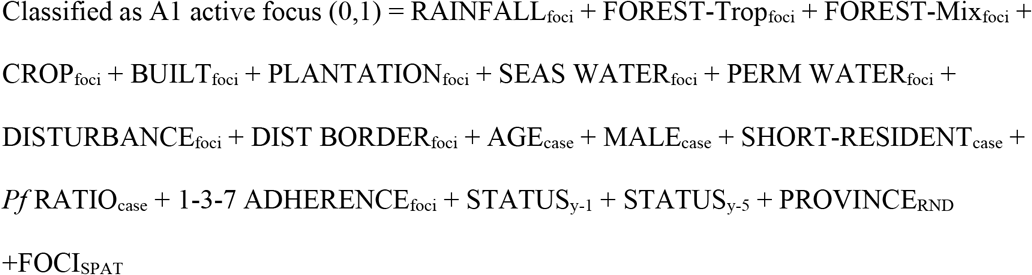

Where:

RAINFALL_foci_: annual monthly mean rainfall

FOREST-Trop_foci_: the percentage of land occupied by tropical forest

FOREST-Mix_foci_: the percentage of land occupied by mixed forest

CROP_foci_: the percentage of land occupied by crops

BUILT_foci_: the percentage of land occupied by built area

PLANTATION_foci_: the percentage of land occupied by plantation area

SEAS WATER_foci_: the percentage of land occupied by seasonal water bodies

PERM WATER_foci_: the percentage of land occupied by permanent water bodies

DISTURBANCE_foci_: the level of forest disturbance

DIST BORDER_foci_: the distance in kilometers from an international border

AGE_case_: the median age of reported cases from FY2016 to FY2020

MALE_case_: the percentage of males among reported cases from FY2016 to FY2020

SHORT-RESIDENT_case_: the percentage of people among reported cases from FY2016 to FY2020 who have lived in Thailand for less than 6 months

*Pf* RATIO_case_: the percentage of *P. falciparum* among reported cases from FY2016 to FY2020

1-3-7 ADHERENCE_foci_: percentage of malaria cases in each focus managed without delays (i.e., adherence to 1-3-7 surveillance protocols in full and on time)

STATUS_y-1_: focus status of the previous year

STATUS_y-5_: the number of years with reported indigenous cases (i.e., A1 focus classification), up to 5 years

PROVINCE_RND_: the focus’ province, included as random effect

FOCI_SPAT_: spatial random effect at the foci level represented by a geospatial spline

Variable selection was performed to identify those factors that have the highest prediction power. The variable selection was based on comparing the Akaike Information Criterion (AIC) of all tested models [26]. Collinearity among the model variables was tested using the variance inflation factor (VIF), which estimates variance inflation due to variable multicollinearity. A high VIF indicates low reliability of a regression model due to strong correlation among its variables, and a VIF >2.5 indicates a level of multicollinearity able to bias its results. The statistical analyses were performed using the R language [27].

## Results

### Changes over time in active foci

The number of active foci for all malaria species in Thailand decreased by 77.7% over the study period, from 1,915 to 427 foci. However, the decline demonstrated an inverse exponential trend, with fewer active foci cleared each year. Between FY2013 and FY2014, the first year of the study, 374 active foci (A1) gained A2 status (19.5%). In contrast, between FY2019 and FY2020, at the study’s endline, only 45 active foci were reclassified to A2 (9.5%) (***Figure 1***).

**Figure 1.**
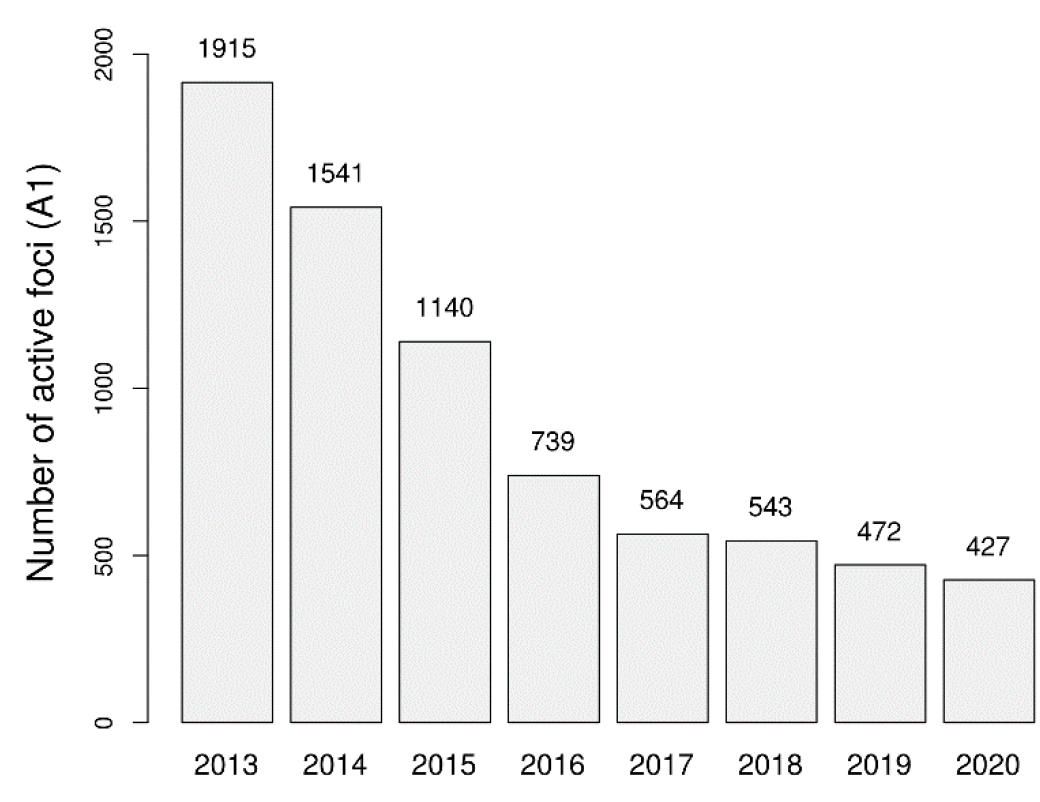
Number of reported active foci from FY2013 to FY2020.

Reported A1 and A2 foci were highly concentrated along international borders, particularly Thailand’s western border with Myanmar (***Figure 2***). Many active foci throughout Thailand gained A2 status during the study period, most notably near the eastern border with Lao PDR (Sakhon Nakhon, Mukdahan, and Ubon Ratchathani provinces) and the northeastern border with Myanmar (Chiang Mai and Mae Hong Son provinces). However, persistent A1 foci (those that retained active foci status consistently for 5 years, from FY2016–FY2020) were almost exclusively found immediately adjacent to borders, showing increasing concentration from A2 to A1 to persistent A1 status (***Figure 2***).

**Figure 2.**
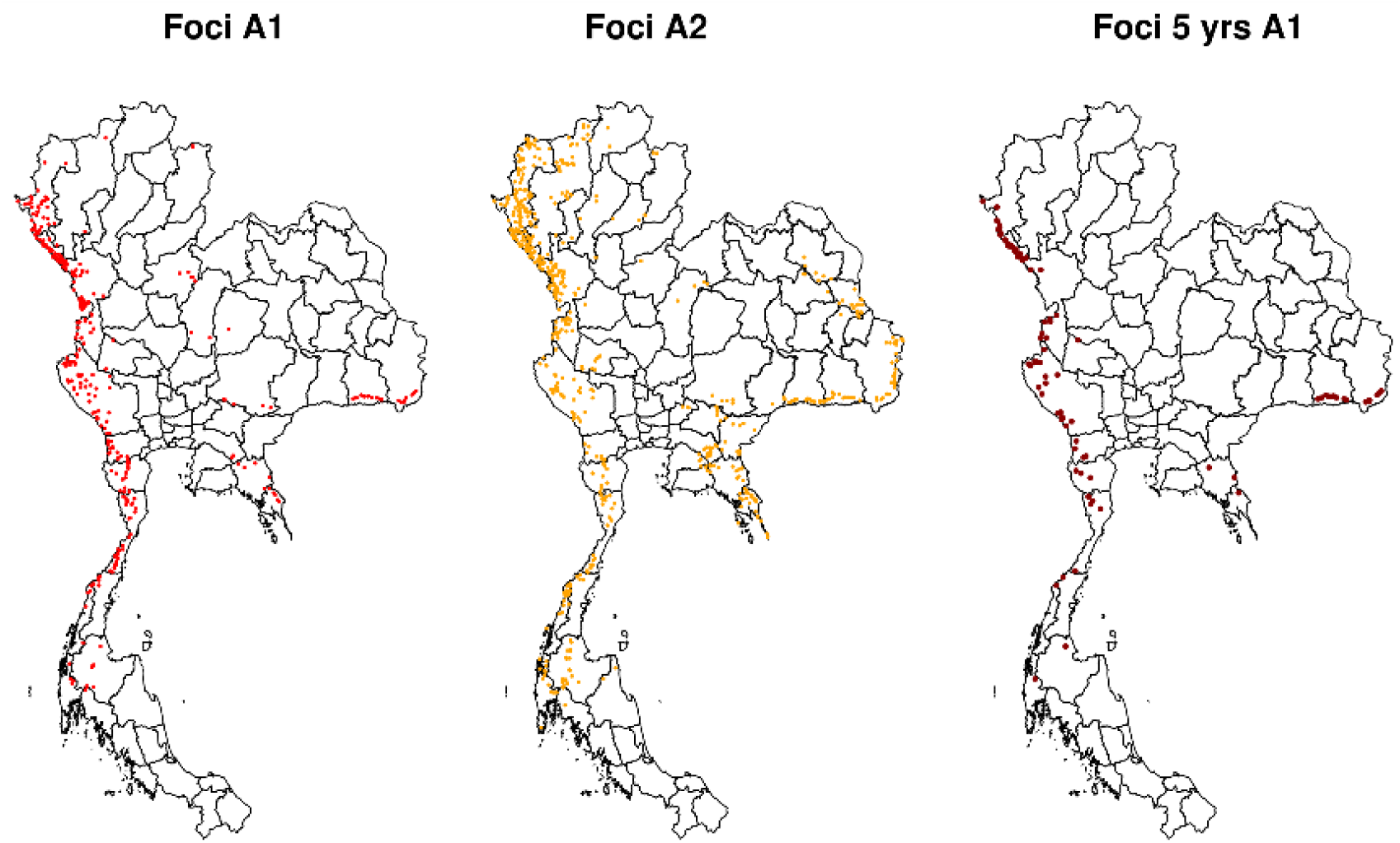
Spatial pattern of A1 and A2 foci and persistent A1 foci (as foci reported as active throughout the time window FY2016 to FY2020)

During the study period, there was an 80.1% decrease in the number of A1 foci reporting a high proportion of *P. falciparum* (80% or higher) among all cases (***Figure S1***). This trend is particularly pronounced along the Cambodian border. It mirrors a national trend of very few foci by FY2020 with greater than 20% of cases represented by *P. falciparum* infections.

### Assessment of environmental composition

A1 foci identified during FY2020 are overlaid on the land cover map, showing that areas with high density of A1 foci are characterized by high forest coverage and located along Thailand’s international borders (***Figure 3***). The land cover map also shows that a high fraction of central Thailand is dedicated to cropland with no active foci (Figure 3).

**Figure 3.**
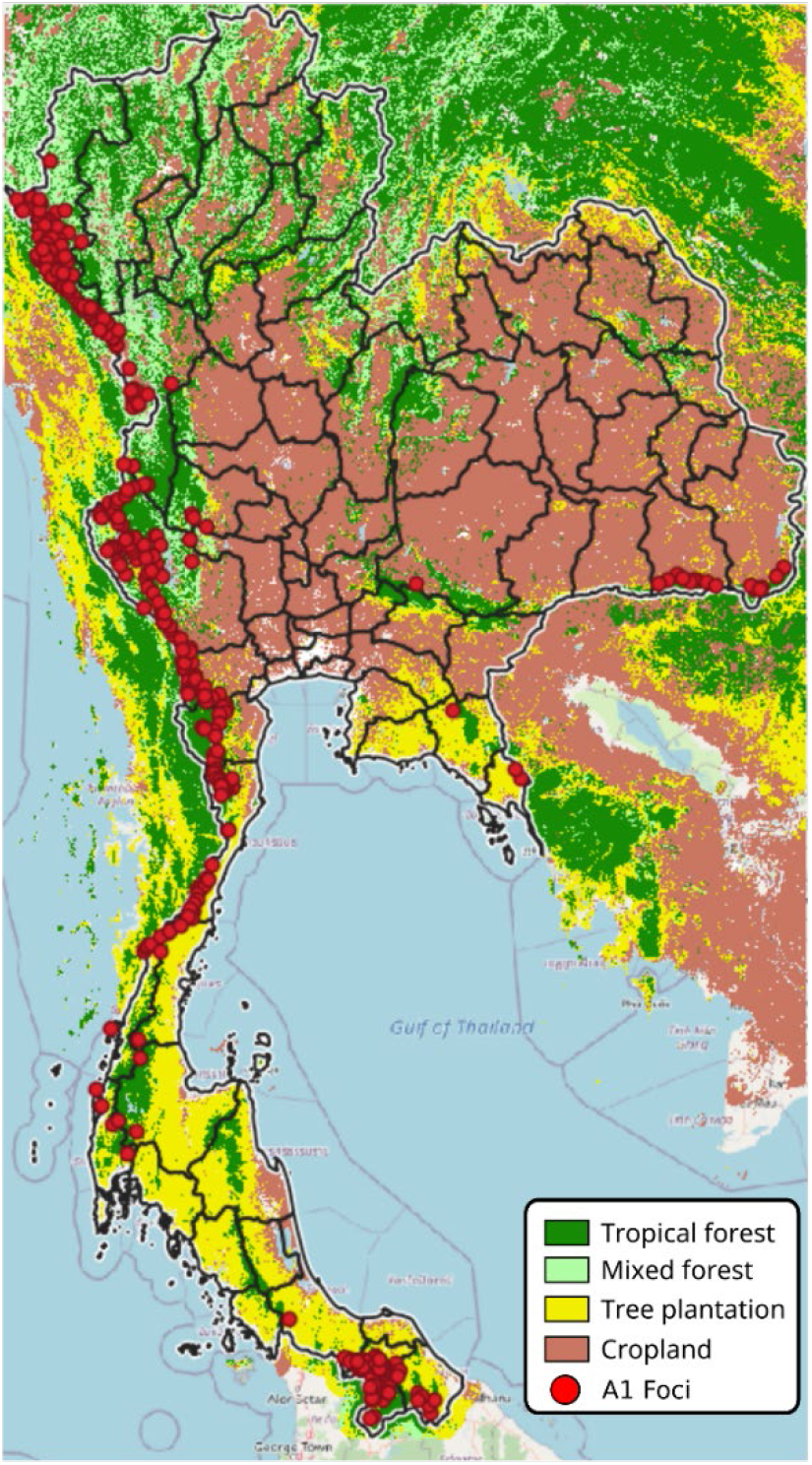
Areas covered by forest, plantation, and cropland with A1 foci, FY2020.

The areas surrounding A1 foci at the borders with Myanmar (A, C), Cambodia (B), and Malaysia (D) are magnified in ***Figure 4***. There is heterogeneity in the habitats surrounding active foci. In Mae Hong Son and Tak provinces in the northwest of Thailand (2A), active foci are surrounded by forest; however, on the Myanmar side of the border, the environment is a mix of forest with plantation. In the other hotspot areas, plantations are notable, and they are usually close to forests. Plantations are also the dominant habitats surrounding A1 foci close to the border with Malaysia (D).

**Figure 4.**
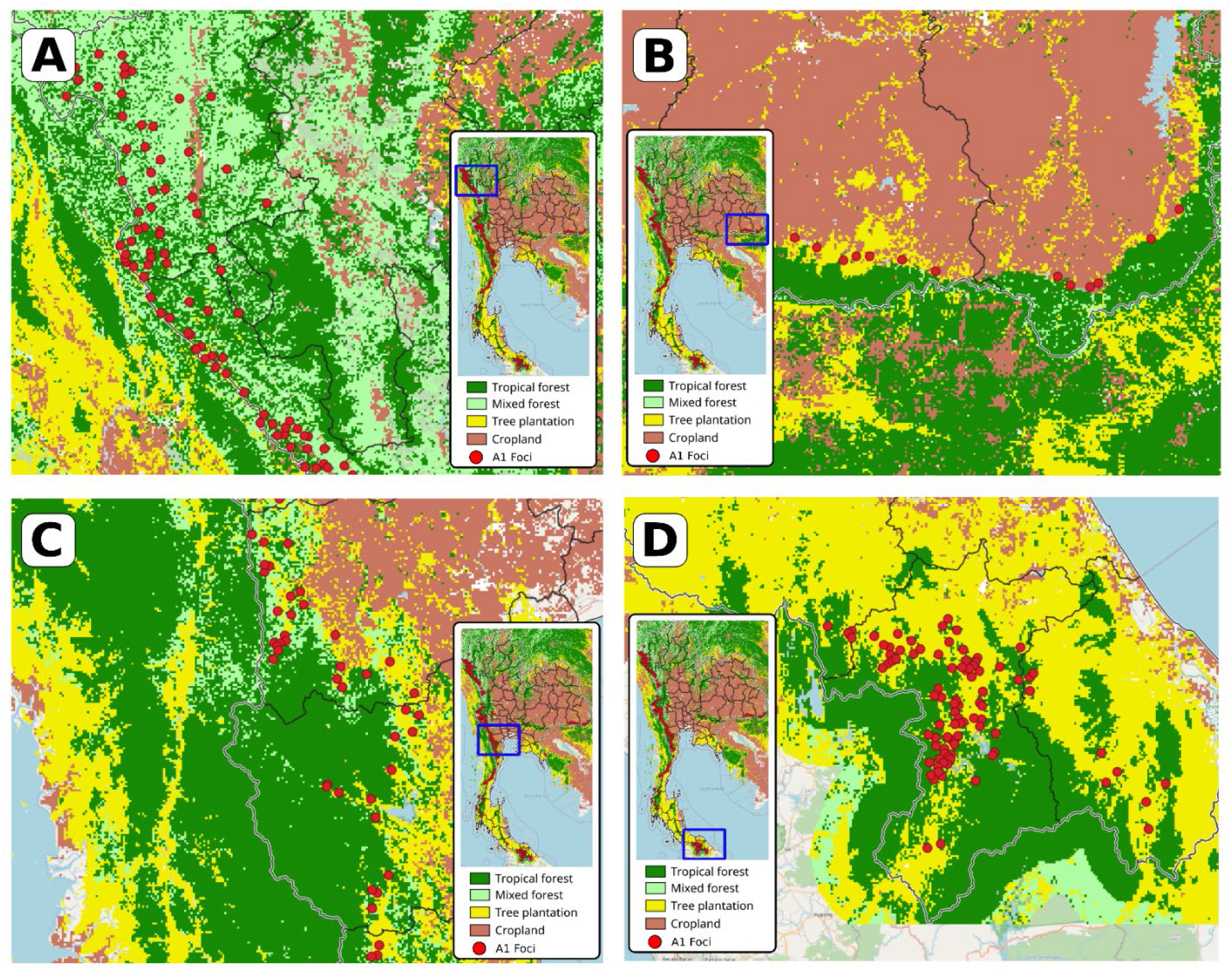
Areas covered by forest, plantation, and cropland around A1 foci in northwestern (A), eastern (B), central (C), and southern (D) Thailand.

The mean percentages of territory covered by each environmental feature in a 1-km radius around the foci are summarized in ***Table 2***. A1 foci showed much higher proportional land cover of plantation, tropical forest, mixed forest, and evergreen forest, in addition to greater forest disturbance. Conversely, A1 foci showed lower proportional urban areas and rice fields. Comparing the environment among the four foci classes showed that land covered by tropical forest and plantation are significantly higher for A1 foci compared to A2, B1, and B2 foci (Kruskal-Wallis, *p*<0.05).

**Table 2.** Environmental characteristics across foci classification. The table shows the mean (among all foci in each classification) percentage of land covered by each environmental feature.

| Focus Classification in FY2020 | Distance from Border (Mean) | Forest Loss Score (Mean) | Fraction Plantation (Mean) | Fraction Tropical Forest (Mean) | Fraction Mixed Forest (Mean) | Fraction Evergreen Forest (Mean) | Fraction Urban Area (Mean) | Fraction Rice Field (Mean) |
| --- | --- | --- | --- | --- | --- | --- | --- | --- |
| A1 | 54.8 km | 14.1 | 20.9% | 10.8% | 14.1% | 1.53% | 0.07% | 0% |
| A2 | 77.85 km | 9.8 | 18.09% | 8.45% | 13.36% | 1.39% | 0.19% | 0% |
| B1 | 119.56 km | 12.39 | 17.33% | 7.22% | 7.40% | 2.30% | 0.61% | 0.32% |
| B2 | 188.69 km | 12.54 | 6.70% | 1.84% | 0.89% | 0.09% | 1.00% | 3.37% |

The annual rainfall trend from FY2015 to FY2020 was very similar across the foci categories, with an increase of rainfall from FY2017 to FY2018, followed by lower precipitation through the study’s endline (***Figure S2***).

### Results of statistical modelling

The multicollinearity analysis indicated a VIF=1.34, showing that model’s results were not affected by multicollinearity. The model selection identified the best fit model as the one including the following variables: FOREST-Trop_foci_, PLANTATION_foci_, DISTURBANCE_foci_, DIST BORDER_foci_, MALE_case_, SHORT-RESIDENT_case_, 1-3-7 ADHERENCE_foci_, STATUS_y-1_, and STATUS_y-5_ (Table 2). This result showed that tropical forest, plantations, forest disturbance, distance from international borders, and historical foci classification are associated with the probability to report indigenous cases, in addition to sex and nationality (***Table 3***). The probability of a focus to be classified as A1 significantly increases when the focus’ environment is mostly composed of tropical forest or plantation, with high forest disturbance. The model also showed that the odds of finding indigenous cases increased for those foci closer to international borders (with Myanmar, Cambodia, and Lao PDR) and that have been reported as A1 during the previous years. A1 classification was also linked to a high fraction of reported cases among males and among short-term resident populations.

**Table 3.**
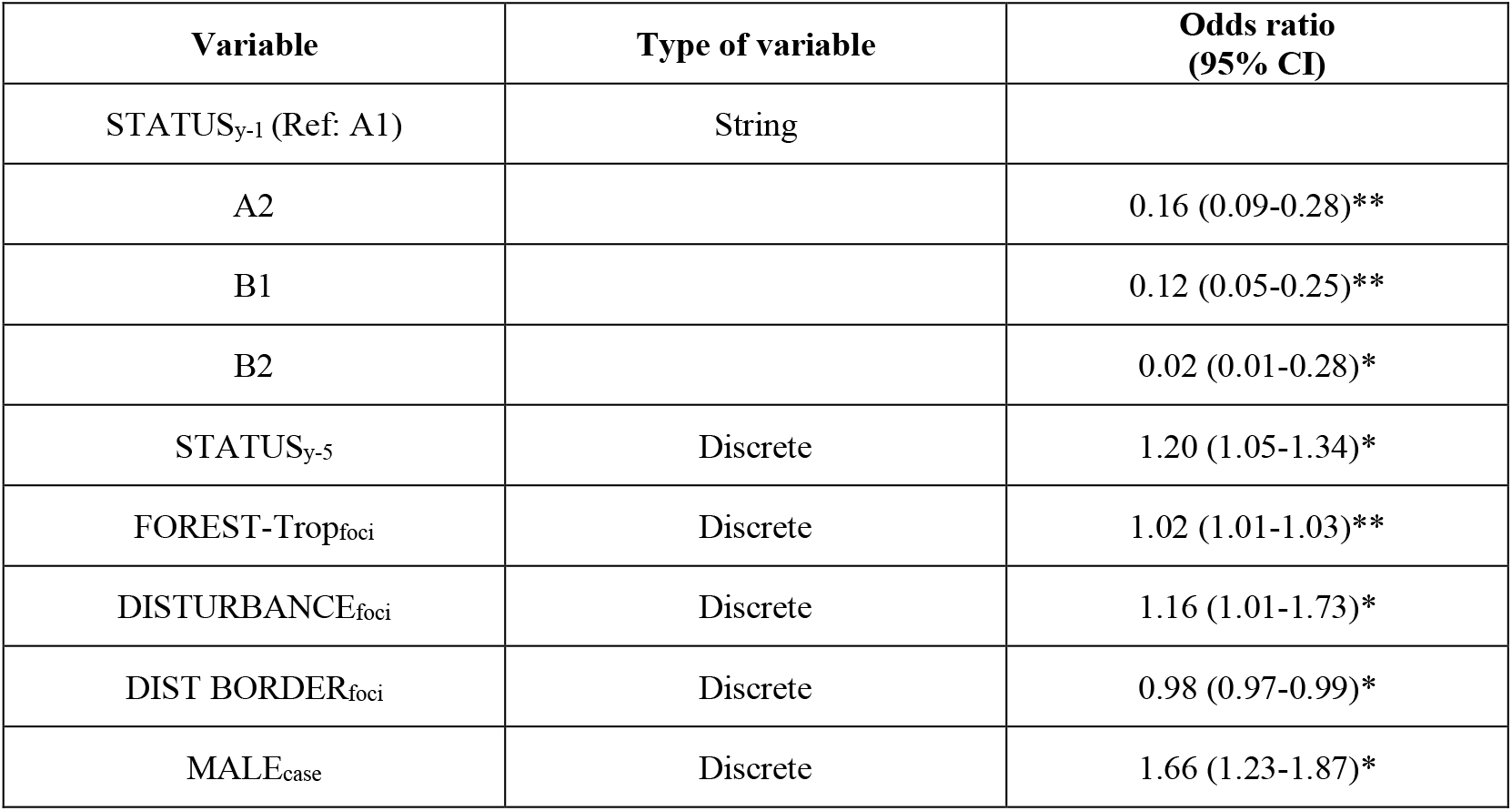
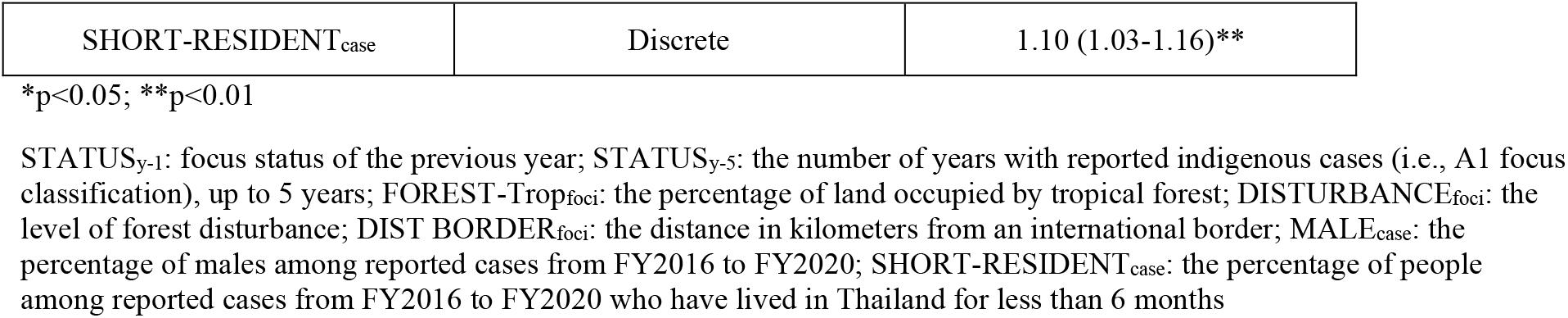
Results of the logistic regression performed with the STAR model.

## Discussion

The study confirms that Thailand’s remaining active foci are clustered along international borders, with a particularly high density along the western border with Myanmar and a high association with tropical forest, plantation, and forest disturbance. These results support the DVBD’s longstanding programmatic focus on border areas and forest-going populations.

Heterogeneous land cover may partially explain these patterns and may contribute to geographically varied success of malaria elimination interventions. The environment of the Thailand-Myanmar border, which includes mountainous regions, forests, and wet-rice fields, is less urbanized than many of Thailand’s low-lying regions [10]. These forested areas provide ideal habitats for *Anopheles* mosquitoes, and access may be more difficult for public health teams to reach for crucial interventions like reactive case detection [28], thereby increasing likelihood of delayed response and onward transmission. By contrast, urbanized or agriculturally developed areas provide a less ideal vector habitat and are more easily accessible [29, 30]. Substantial prior research from malaria-endemic countries suggests that decreased risks of infection are correlated with urbanization [31, 32].

Other environmental factors, including distance from seasonal or permanent bodies of water and rainfall, have previously been shown to impact malaria transmission [33], but were not significant predictors in our model. Temporal variations (wet vs. dry season), rather than geographic differences, in rainfall distribution may be responsible for these results.

However, the study also showed a substantial, yet slowing, decrease in the number of active malaria foci in Thailand since FY2013. These results are consistent with the 2021 World Malaria Report, which found that global progress against malaria had reached a plateau before the start of the ongoing coronavirus disease 2019 (COVID-19) pandemic [34]. However, these slowing rates indicate that additional or revised surveillance efforts, coupled with other new interventions, will be necessary to meet the elimination goals.

Thailand’s routine surveillance data show that active foci with significantly lower adherence to 1-3-7 protocols are in border areas with these environmental factors, but which also experience additional challenges [11]. These challenges include waning drug efficacy, migration linked with regional sociopolitics, and civil unrest. To continue to accelerate to elimination, Thailand may need to further refine the 1-3-7 approach to address persisting challenges in adherence, perhaps enhancing protocols to better suit these specific micro-contexts, although developing such protocols would require additional resources. The DVBD may also need to consider more aggressive strategies such as targeted chemoprevention, a revised active case detection program to target emerging high-risk populations, enhanced follow-up schemes or new treatments like tafenoquine for the radical cure of *P. vivax*, support for improved data recording and use, and involvement of other actors such as the private sector and military to improve reach of malaria interventions.

The results also indicated that demographic factors, such as percentage of males and short-term resident cases, increase the probability of a focus to report a local case. Successfully eliminating malaria will largely depend on targeting specific population groups to ensure high coverage of essential interventions. In Thailand, men between 15 and 44 years of age are more likely to work and sleep overnight in forests; they also comprise 65.9% of malaria infections [35]. Thailand is the principal recipient country of cross-border workers in the GMS [36], and population movement is influenced by social and economic crises including COVID-19 and the 2021 political crisis in Myanmar. These communities often live in hard-to-reach areas and may face more barriers to accessing malaria care than longer-term residents of Thailand [37]. The DVBD will need to find safe and flexible ways to meet their goal of providing effective and complete malaria treatment to all patients in Thailand. Continuing to focus on specific areas with substantial transboundary travel and migration could reduce local transmission.

Environmental characteristics overlap to affect vector reproduction, exposure risk, and access to malaria health services, thereby linking environmental and social patterns [10]. For example, a highly forested area may provide outdoor work opportunities, in turn increasing exposure to exophagic mosquitoes [9, 15, 37]. The results of this analysis suggest that environmental factors alone are not driving malaria transmission in Thailand but rather a combination of syndemic factors. Human activities in areas covered by tropical forests and plantations may result in malaria importation and in turn, local transmission.

These results, which highlight areas at high risk of reporting indigenous cases, are already being used in the DVBD’s risk planning to prevent reestablishment in malaria-free areas. As active foci continue to be eliminated, the DVBD will need to remain vigilant against the return of active foci in susceptible areas. As documented in Sudathip (2021), movement between A1 and A2 status is frequent, which may be unsurprising due to Thailand’s sensitive foci classification criteria: having just one indigenous case reverts a focus from A2 to A1 status [5]. The DVBD plans to develop an automated stratification that will maximize use of Thailand’s existing environmental, climate, and demographic data with minimal statistical input. This stratification will be programmed into an open-source software that enables interactive maps, so that district malaria officers across Thailand will be able to review their jurisdiction’s results and corresponding planning and response actions. The DVBD itself will manage data sources and anticipated update cycles, maintenance of current data in the software, and analysis and use from the resulting maps and tables. There may also be something to learn from provinces or foci where malaria has been reintroduced, some of which are not on international borders. The stratification will employ a non-modeling method to categorize subdistricts into malaria transmission strata, with corresponding POR interventions for each stratum. The DVBD will also issue guidance by stratum for response teams to use a reference for what POR actions need to be maintained and what actions would be taken in the case of an unexpected case or outbreak.

## Conclusion

This analysis suggests that environmental factors alone are not driving malaria transmission in Thailand; rather, other factors, including demographics and behaviors, may be contributors. However, these factors are syndemic, so human activities in areas covered by tropical forests and plantations may result in malaria importation that could lead to local transmission. The environmental and demographic determinants of malaria transmission identified in this study support greater understanding of the vulnerability and receptivity bases of POR planning. The DVBD will continue to tailor its surveillance and response to protect high-risk populations and environments and to prevent malaria reintroduction and/or reestablishment. Although these findings are specific to Thailand, this study reveals a generalizable link between both environmental conditions and human activities with malaria elimination. This type of analysis may help other countries adapt their malaria elimination and POR programming to promote consistent progress.

## Data Availability

The visualizations supporting the conclusions of this article are available in the Malaria Online repository.

http://malaria.ddc.moph.go.th/.

## List of abbreviations

COVID-19: coronavirus disease 2019
DVBD: Division of Vector Borne Diseases
FY: fiscal year
GMS: Greater Mekong Subregion
Lao PDR: Lao People’s Democratic Republic
MIS: Malaria Information System
MOPH: Ministry of Public Health
NMES: National Malaria Elimination Strategy
PMI: President’s Malaria Initiative
POR: prevention of reestablishment
USAID: United States Agency for International Development
WHO: World Health Organization

## Declarations

### Ethics approval and consent to participate

Not applicable.

### Consent for publication

Not applicable.

### Availability of data and materials

The visualizations supporting the conclusions of this article are available in the Malaria Online repository, http://malaria.ddc.moph.go.th/.

### Authors’ contributions

PS, DB, and JAS conceptualized the manuscript. DB and AP organized and cleaned the data. DB analyzed the spatial results and interpreted the results with support from JAS. IP led the writing of the manuscript with support from DB and JAS. All authors, including PP, JK, DG, CS, NP, and DS, provided input and review. All authors read and approved the final manuscript.

## Acknowlegments

The authors recognize all local public health teams and partners who have contributed to the malaria elimination program at the national and subnational levels under the leadership of Thailand’s Division of Vector Borne Diseases, Department of Disease Control, Ministry of Public Health.

## Authors’ information

The contents of this article are the responsibility of the authors and do not necessarily reflect the views of USAID, PMI, or the U.S. Government. DG is a staff member of the World Health Organization (WHO) and is responsible for the views expressed in this publication, which do not necessarily reflect the decisions or policies of the WHO.

## Appendix

**Figure S1.**
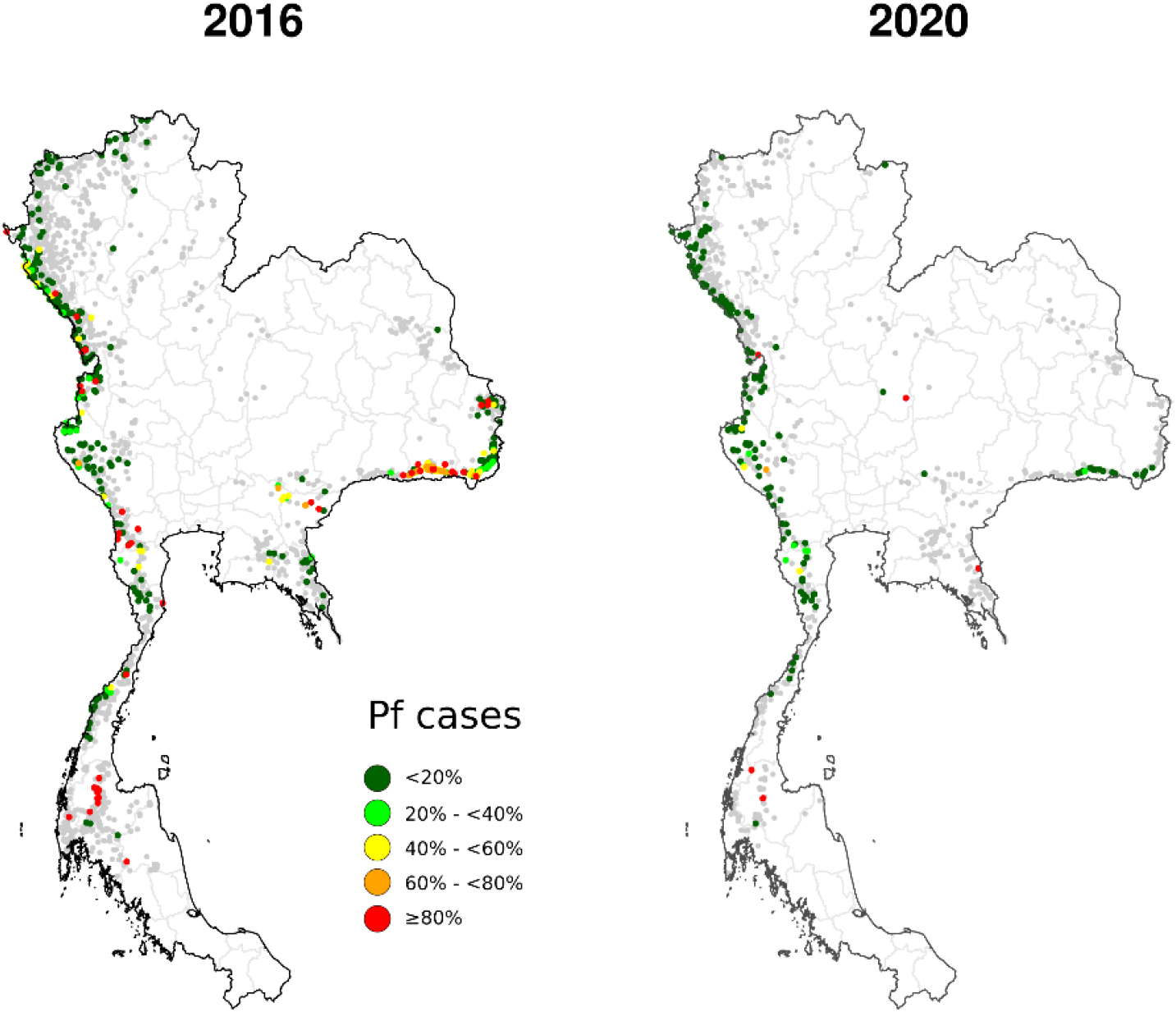
Percentage of *Pf* cases among reported cases in A1 foci in FY2016 and FY2020.

**Figure S2.**
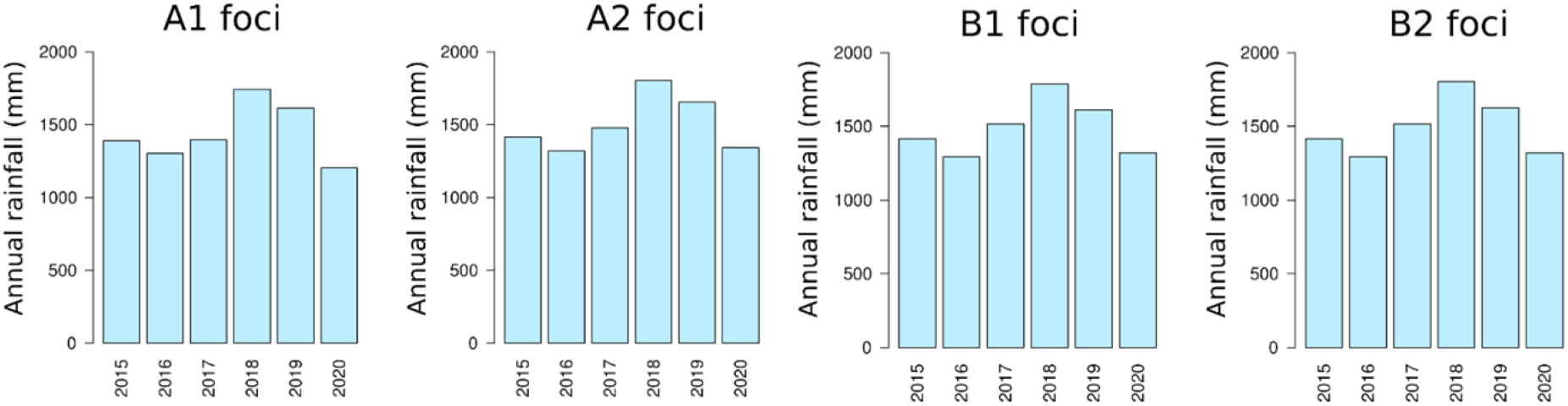
Median annual rainfall per foci classification, FY2015–FY2020.

